# Non-occupational and occupational factors associated with specific SARS-CoV-2 antibodies among Hospital Workers – a multicentre cross-sectional study

**DOI:** 10.1101/2020.11.10.20229005

**Authors:** Christian R. Kahlert, Raphael Persi, Sabine Güsewell, Thomas Egger, Onicio B. Leal-Neto, Johannes Sumer, Domenica Flury, Angela Brucher, Eva Lemmenmeier, J. Carsten Möller, Philip Rieder, Reto Stocker, Danielle Vuichard-Gysin, Benedikt Wiggli, Werner C. Albrich, Baharak Babouee Flury, Ulrike Besold, Jan Fehr, Stefan P. Kuster, Allison McGeer, Lorenz Risch, Matthias Schlegel, Andrée Friedl, Pietro Vernazza, Philipp Kohler

**Affiliations:** Cantonal Hospital St Gallen, Division of Infectious Diseases and Hospital Epidemiology, St Gallen, Switzerland; Children’s Hospital of Eastern Switzerland, Department of Infectious Diseases and Hospital Epidemiology, St. Gallen, Switzerland; Clinical Trials Unit, Cantonal Hospital of St. Gallen, St. Gallen, Switzerland; Epitrack, Recife, Brazil; Department of Economics, University of Zurich, Zurich, Switzerland; Psychiatry Services of the Canton of St. Gallen (South), Switzerland; Clienia Littenheid AG, Private Clinic for Psychiatry and Psychotherapy, Littenheid, Switzerland; Center for Neurological Rehabilitation, Zihlschlacht, Switzerland; Hirslanden Clinic, Zurich, Switzerland; Thurgau Hospital Group, Division of Infectious Diseases and Hospital Epidemiology, Muensterlingen, Switzerland; Swiss National Center for Infection Prevention (Swissnoso), Berne, Switzerland; Kantonsspital Baden, Division of Infectious Diseases and Hospital Epidemiology, Baden, Switzerland; Geriatric Clinic St. Gallen, St. Gallen, Switzerland; Department of Public and Global Health, University of Zurich, Zurich, Switzerland; Federal Office of Public Health, Bern, Switzerland; University Hospital and University of Zurich, Division of Infectious Diseases and Hospital Epidemiology, Zurich, Switzerland; Sinai Health System, Toronto, Canada; Labormedizinisches Zentrum Dr Risch Ostschweiz AG, Buchs, Switzerland; Private Universität im Fürstentum Liechtenstein, Triesen, Liechtenstein; Center of Laboratory Medicine, University Institute of Clinical Chemistry, University of Bern, Inselspital, Bern, Switzerland

**Keywords:** COVID-19, Seroprevalence, Healthcare workers, Switzerland, Risk factors

## Abstract

**Objectives:** Protecting healthcare workers (HCW) from Coronavirus Disease-19 (COVID-19) is critical to preserve the functioning of healthcare systems. We therefore assessed seroprevalence and identified risk factors for Severe Acute Respiratory Syndrome-Coronavirus-2 (SARS-CoV-2) seropositivity in this population.

**Methods:** Between June 22^nd^ and August 15^th^ 2020, HCW from institutions in Northern/Eastern Switzerland were screened for SARS-CoV-2 antibodies. We recorded baseline characteristics, non-occupational and occupational risk factors. We used pairwise tests of associations and multivariable logistic regression to identify factors associated with seropositivity.

**Results:** Among 4’664 HCW from 23 healthcare facilities, 139 (3%) were seropositive. Non-occupational exposures independently associated with seropositivity were contact with a COVID-19-positive household (adjusted OR=54, 95%-CI: 31-97) and stay in a COVID-19 hotspot (aOR=2.2, 95%-CI: 1.1-3.9). Blood group 0 vs. non-0 (aOR=0.4, 95%-CI: 0.3-0.7), active smoking (aOR=0.5, 95%-CI: 0.3-0.9) and living with children <12 years (aOR=0.3, 95%-CI: 0.2-0.6) were associated with decreased risk. Occupational risk factors were close contact to COVID-19 patients (aOR=2.8, 95%-CI: 1.5-5.5), exposure to COVID-19-positive co-workers (aOR=2.0, 95%-CI: 1.2-3.1), poor knowledge of standard hygiene precautions (aOR=2.0, 95%-CI: 1.3-3.2), and frequent visits to the hospital canteen (aOR=1.9, 95%-CI: 1.2-3.1).

**Conclusions:** Living with COVID-19-positive households showed by far the strongest association with SARS-CoV-2 seropositivity. We identified several potentially modifiable risk factors, which might allow mitigation of the COVID-19 risk among HCW. The lower risk among those living with children, even after correction for multiple confounders, is remarkable and merits further study.

## INTRODUCTION

Coronavirus disease 2019 (COVID-19) is currently afflicting healthcare systems around the globe. As of December 2^nd^ 2020, over 1.5 million COVID-19 deaths have been reported worldwide [1]. In Switzerland, over 350’000 COVID-19 cases have been reported, more than 14’000 patients have been hospitalized and over 5’000 have died [2]. Seroprevalence studies among Swiss healthcare workers (HCW) performed in March and April 2020 have shown a low prevalence of 1% in the Eastern part of the country, and a higher prevalence of around 10% in the Western part [3,4]. The recent massive re-emergence of cases in many European countries including Switzerland is putting further strain on healthcare systems and hospital workers. Studies from different countries suggest that HCW are at increased risk to acquire Severe Acute Respiratory Syndrome-Coronavirus-2 (SARS-CoV-2) when compared to the general population [5]. In the UK, HCW and their household contacts accounted for a sixth of all COVID-19 cases admitted to the hospital for those aged 18-65 years. This risk was increased for HCW involved in patient care [6]. In light of these data it is imperative to better understand risk factors for SARS-CoV-2 acquisition among HCW in order to better protect them from infection.

In this multicentre study from Switzerland, we aimed to assess the prevalence of specific antibodies against SARS-CoV-2 among HCW with and without patient contact. In addition, we identified non-occupational and occupational factors associated with seropositivity to inform prevention recommendations for this population.

## METHODS

### Study design and participants

We initiated a multicentre cross-sectional study between June 22^nd^ and August 15^th^ 2020 in healthcare institutions located in Northern and Eastern Switzerland. Acute care hospitals, rehabilitation clinics, geriatric and psychiatric clinics in the region were asked to participate. Within every participating institution, employees aged 16 years or older were invited to enrol into the study via institutional webpages. Employees registered online and provided electronic consent. The study was approved by the ethics committee of Eastern Switzerland (#2020-00502).

### Questionnaire and definitions

We implemented a multi-modular digital web-based questionnaire for institutions and participants. Questions about facility structure were asked in the institutional questionnaire. Participants received an invitation to the questionnaire by email and were asked about anthropometric data, occupational and non-occupational risk exposures, and previous SARS-CoV-2 nasopharyngeal swabs. Household contacts were defined as people living in the same household or intimate partners; close contact to COVID-19 patients was assumed for those with contact >15 minutes within 2 meters with or without personal protective measures (PPE); aerosol-generating procedures (AGP) were defined according to guidelines of the Swiss Center for Infection Prevention (Swissnoso). Poor knowledge of standard precautions was assumed for those who correctly identified less than 3 measures in a multiple choice question (among a choice of hand hygiene, surgical mask in case of respiratory symptoms, donning gowns in case of potential contamination with body fluids, cough etiquette, and vaccination). Low protection while caring for COVID-19 patients was assumed for those using less than 3 measures out of face masks, gloves, gowns, and goggles.

### Sample processing

Upon registration, participants provided a venous blood sample, which was collected at local sites. Samples were analysed with an electro-chemiluminescence immunoassay (ECLIA, Roche Diagnostics, Rotkreuz, Switzerland, detection of total antibodies directed against the nucleocapsid-(N)-protein of SARS-CoV-2) run on a COBAS 6000 instrument, as described elsewhere [7]. A subgroup of samples with a positive signal in the ECLIA (at a cut-off index, COI, ≥ 1) were also tested with an Enzyme-linked Immunosorbent Assay (ELISA, Euroimmune, Germany, detection each of IgG and IgA antibodies against S1 domain of the spike-(S)-protein including the immunologically relevant receptor binding domain). Cut-offs for seropositivity were applied as recommended by the manufacturers. Seropositivity was defined as positive result in the ECLIA followed by confirmation in the ELISA (either positive IgA or IgG).

### Statistical analysis

The relative frequency of participants with ELISA confirmed positive and negative serology was compared between levels of baseline characteristics, non-occupational risk factors and occupational risk factors. Fisher’s exact test was used for dichotomous factors or factors with a reference level, comparing each level to the reference. Individuals with missing data were removed from the analysis of the respective variable. Logistic regression was used for numeric and ordinal variables. Age, sex, body mass index (BMI), smoking status, comorbidities as well as non-occupational and occupational risk factors expected to influence seropositivity were entered into a multivariable logistic regression model. For sensitivity analysis, we fitted two additional models including place of residence (7 predefined regions) and institution either as fixed effects or as random effects to assess whether spatial proximity or clustering of observations confounded the effects of the risk factors. Analyses were performed with R statistical software, version 4.0.2.

## RESULTS

### Baseline characteristics

We included 17 institutions on 23 sites across Northern and Eastern Switzerland, thereof 19 inpatient sites (14 acute care; 1 geriatric clinic; 1 rehabilitation clinic; 3 psychiatric clinics) and 4 outpatient clinics (3 psychiatric facilities; 1 blood donation centre). The total of represented patient beds was 3’523 (thereof 106 ICU beds) (**Table 1**).

**Table 1.** Characteristics of institutions (n=17) including size, number of study participants and seropositivity.

|  | Sites<br>(n) | Inpatients<br>(yes vs no) | Beds<br>(n) | ICU beds<br>(n) | HCW<br>(n) | HCW in<br>study (n) | HCW in<br>study (%) | Seropositive<br>HCW (n) | Seropositive<br>HCW (%) |
| --- | --- | --- | --- | --- | --- | --- | --- | --- | --- |
| TOTAL | 23 | NA | 3'523 | 106 | 17'060 | 4664 | 27% | 139 | 3.0% |
| Acute care | 3 | yes | 765 | 36 | 5930 | 1074 | 18% | 37 | 3.4% |
| Acute care | 1 | yes | 370 | 10 | 2245 | 1023 | 46% | 39 | 3.8% |
| Acute care | 3 | yes | 304 | 7 | 1367 | 534 | 39% | 9 | 1.7% |
| Acute care | 1 | yes | 74 | 0 | 362 | 109 | 30% | 3 | 2.8% |
| Acute care | 1 | yes | 46 | 0 | 178 | 66 | 37% | 1 | 1.5% |
| Acute care | 1 | yes | 246 | 9 | 749 | 169 | 23% | 7 | 4.1% |
| Acute care | 1 | yes | 310 | 12 | 740 | 171 | 23% | 3 | 1.8% |
| Acute care | 1 | yes | 330 | 18 | 1788 | 448 | 25% | 18 | 4.0% |
| Acute care | 1 | yes | 129 | 6 | 525 | 159 | 30% | 4 | 2.5% |
| Acute care | 1 | yes | 100 | 8 | 632 | 109 | 17% | 3 | 2.8% |
| Geriatric acute<br>care | 1 | yes | 98 | 0 | 265 | 123 | 46% | 3 | 2.4% |
| Rehabilitation<br>clinic | 1 | yes | 135 | 0 | 510 | 168 | 33% | 7 | 4.2% |
| Psychiatric clinic | 1 | yes | 242 | 0 | 360 | 190 | 53% | 1 | 0.5% |
| Psychiatric clinic | 1 | yes | 150 | 0 | 391 | 108 | 28% | 1 | 0.9% |
| Psychiatric clinic | 1 | yes | 224 | 0 | 780 | 98 | 13% | 1 | 1.0% |
| Psychiatry | 3 | no | NA | NA | 178 | 88 | 49% | 2 | 2.3% |
| Blood donation | 1 | no | NA | NA | 60 | 27 | 45% | 0 | 0.0% |

Abbreviations: ICU, Intensive Care Unit; HCW, Healthcare Workers

Of the 17’060 potentially eligible HCW, 4’664 (27%) participated in the study. Median age was 38 years (range 16-73); 3’654 (78%) were female. The majority were nurses (n=2’126; 46%) followed by physicians (n=776; 17%); 3’676 (79%) reported having patient contact (**Table 2**).

**Table 2.**
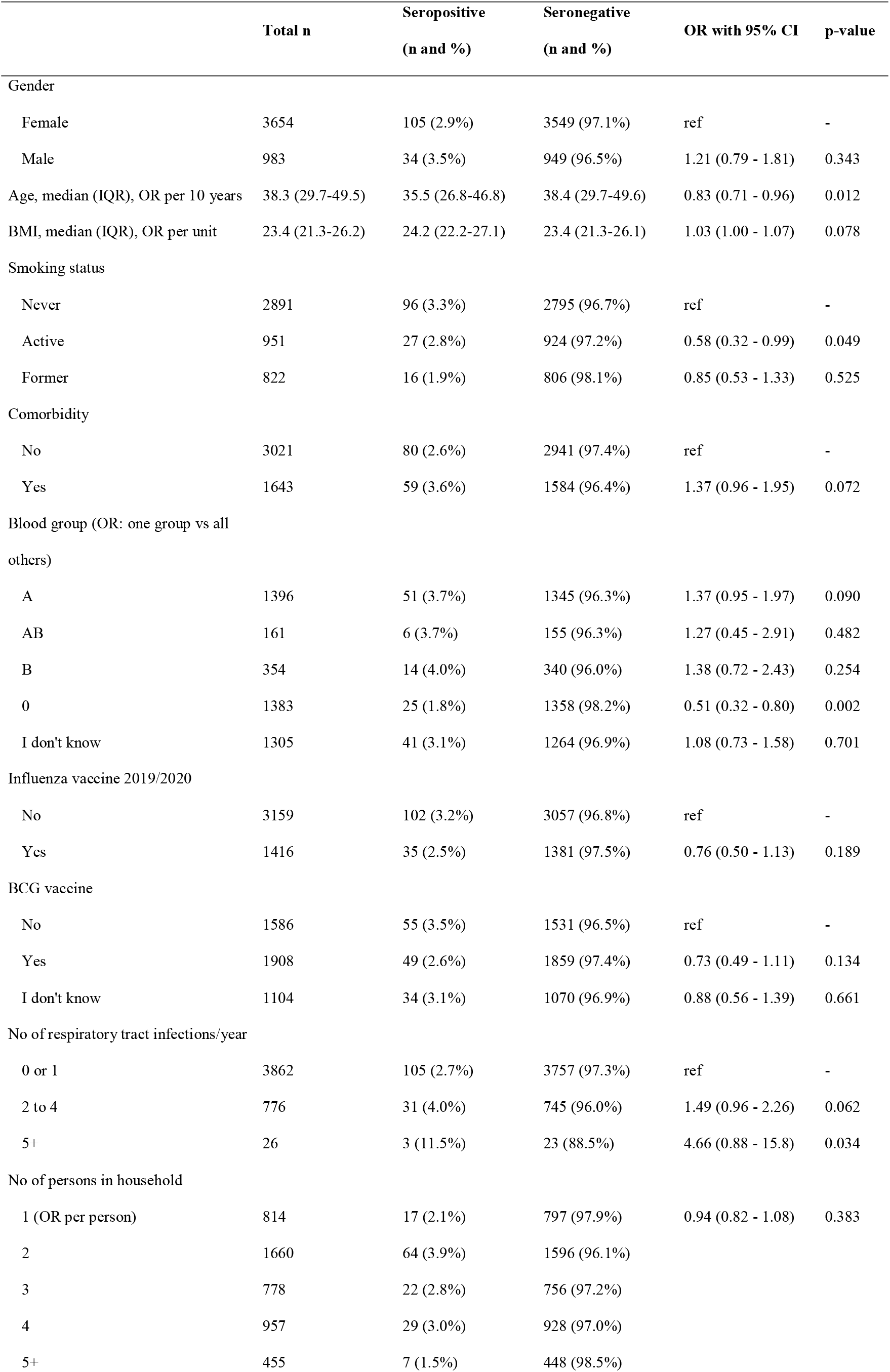
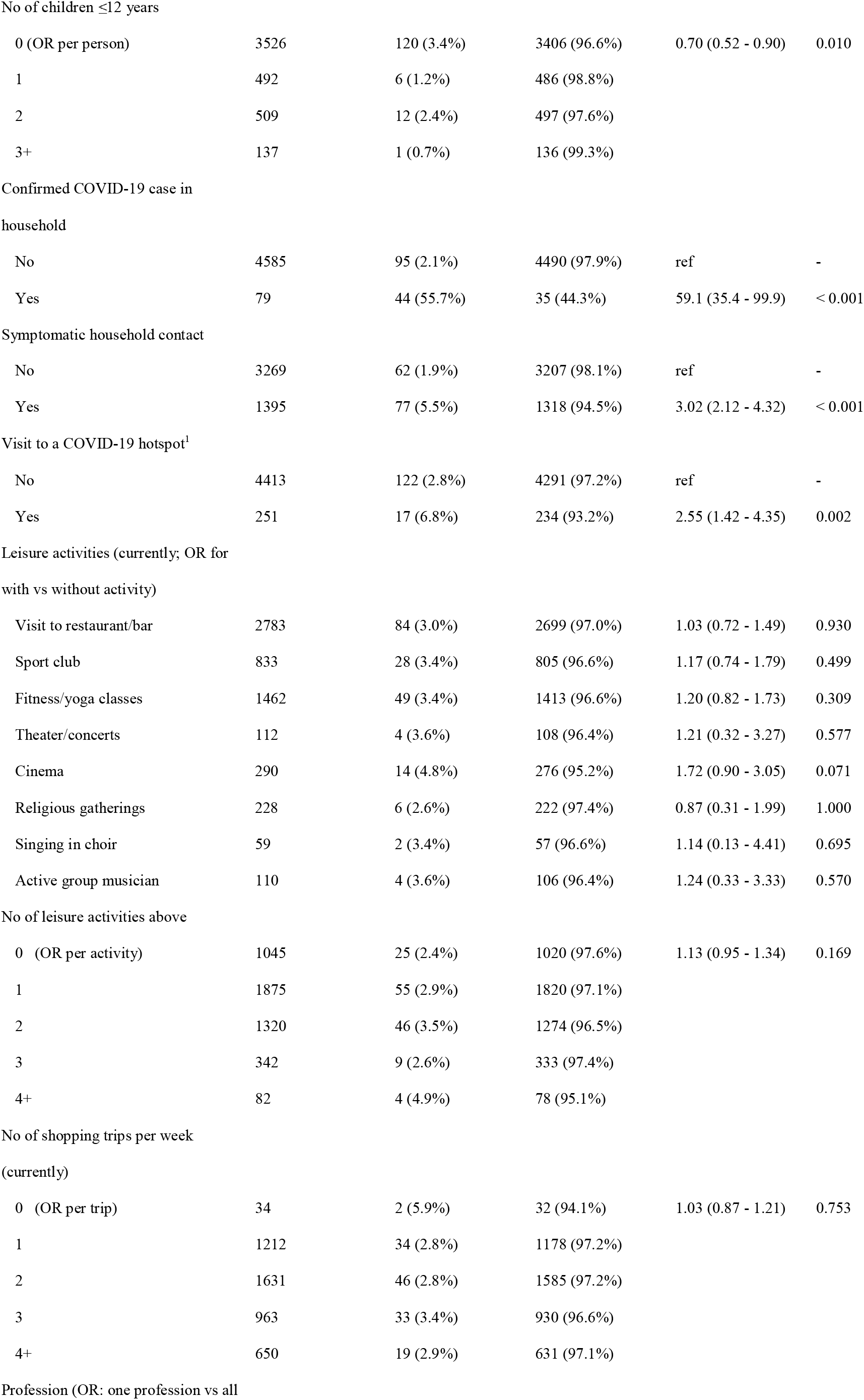
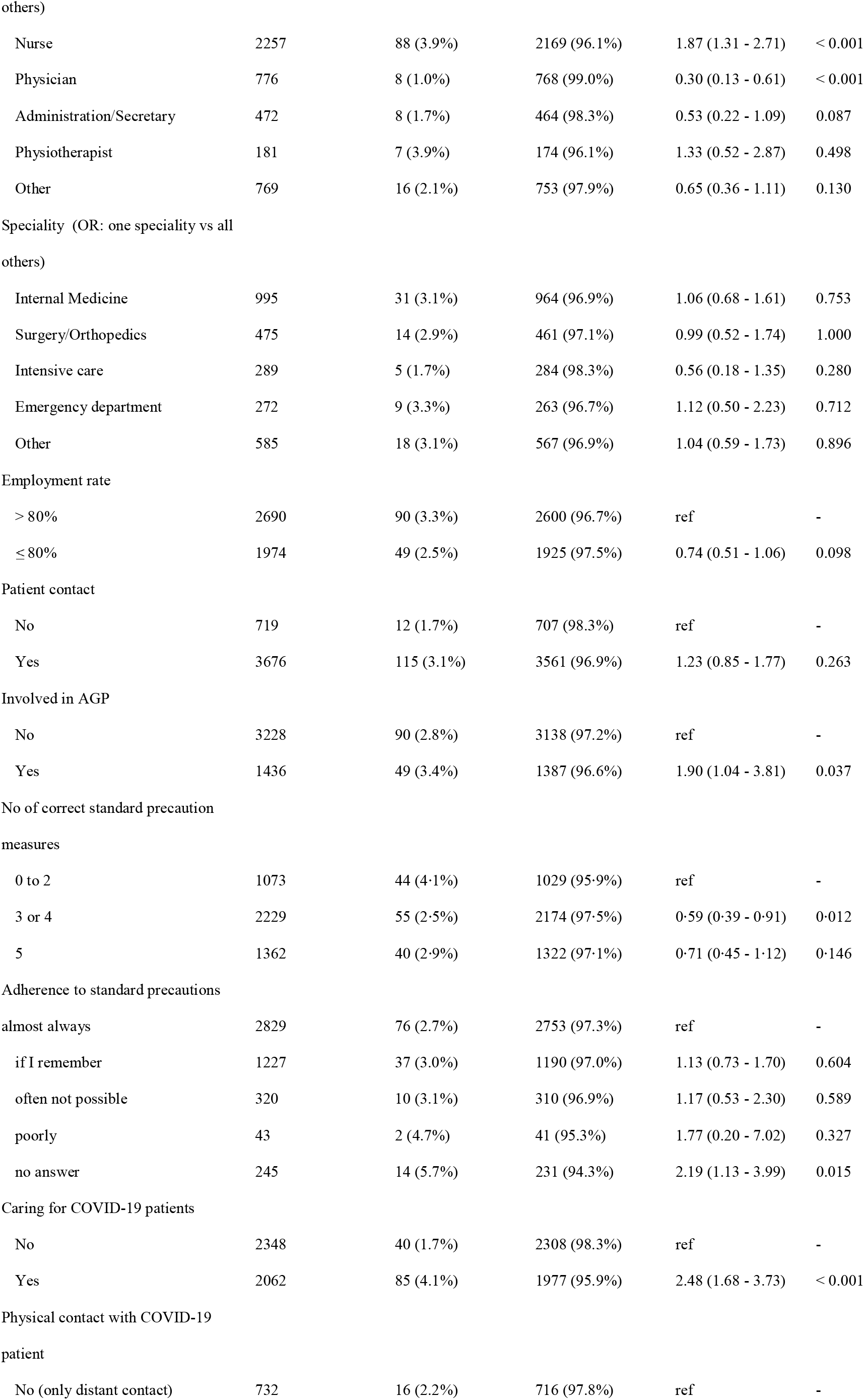
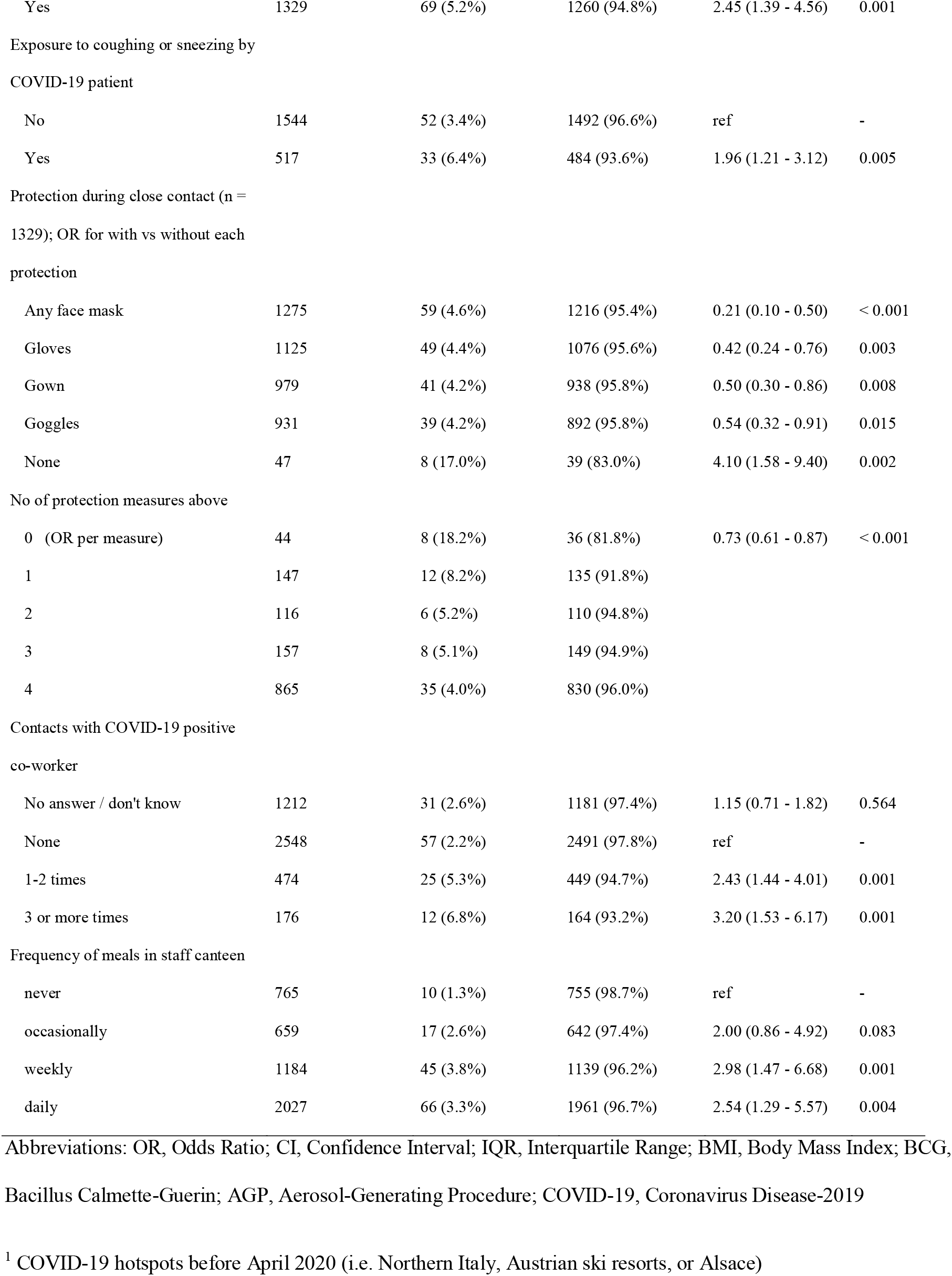
Baseline, non-occupational and occupational factors by serostatus.

### Seropositivity and self-reported PCR results

Overall, seropositivity was 3.0% (139/4’664). Among these 139, 88 (63%) were also tested with the confirmatory ELISA and all 88 samples had either positive IgA or IgG. On the institutional level, seropositivity ranged between 0.5% and 4.2% for inpatient, and 0% and 2.3% for outpatient facilities (**Table 1**). Seropositivity by district ranged from 0% to 13%. Seropositivity was lower in regions located in Eastern compared to Northern Switzerland (**Figure 1**).

**Figure 1.**
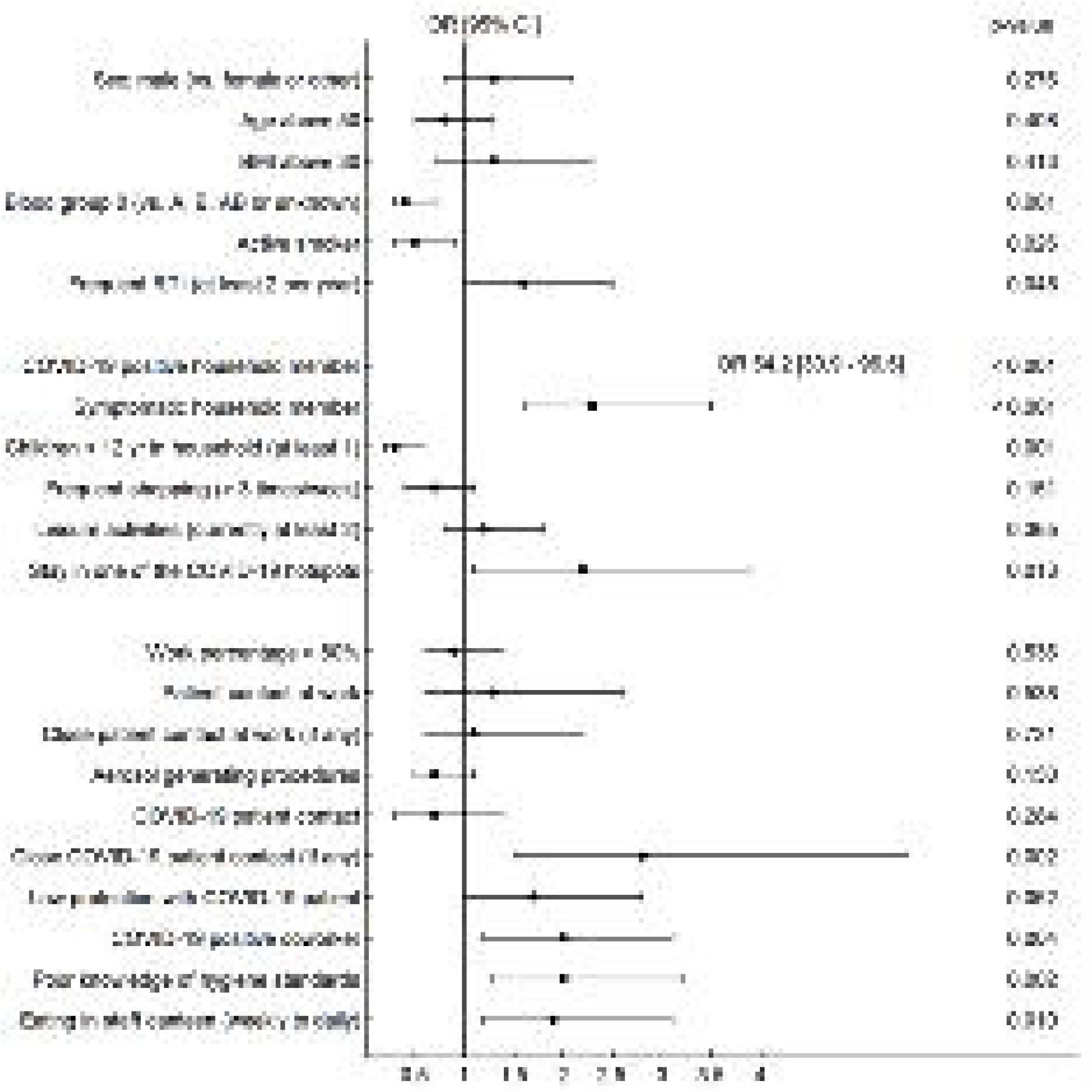
SARS-CoV-2 seropositivity by district (place of residence of healthcare workers) in Northern and Eastern Switzerland (in grey: no seroprevalence indicated for districts with less than 10 participants).

A previous PCR result was reported by 864 of 4’664 (18.5%) participants. Of the 72 participants with positive PCR, 66 (92%) were also seropositive. On the other hand, 17/792 (2.2%) participants with negative PCR had a positive serology. Overall, 23/864 (2.7%) self-reported PCR results were discordant to serology results. Seroprevalence among those without previous PCR was 1.5% (56/3’800).

### Non-occupational factors associated with seropositivity

Exposure to COVID-19 confirmed (55.7% vs. 2.1%, p<0.001) or symptomatic, not confirmed household contacts (5.5% vs. 1.9%, p<0.001) was strongly associated with seropositivity. Also, having visited a known COVID-19 hotspot in Austria (but not in Italy or France) was clearly associated with seropositivity (6.8% vs. 2.8%, p=0.002). Seroprevalence was lower among those with blood group 0 vs. non-0 (1.8% vs. 3.5%, p=0.002) and for those living with children aged 12 or younger (1.7% vs. 3.4%, p=0.002) (**Table 2**).

### Occupational factors associated with seropositivity

Nurses had a higher (3.9%), physicians a lower (1.0%) seropositivity rate; no differences according to medical speciality were noted. Seroprevalence was higher among those with patient contact (3.1% vs. 1.7%, p=0.037), particularly for those with contact to confirmed COVID-19 patients (4.1% vs. 1.7%, p<0.001). Workers indicating low protection while caring for COVID-19 patients (5.8% vs. 3.5%, p=0.019) and those with poor knowledge of hygiene standards had higher seropositivity (4.1% vs. 2.6%, p=0.018) (**Figure 2, panels A/B**). The number of unprotected contacts to COVID-19 confirmed or symptomatic co-workers was associated with seropositivity (**Figure 2, panel C**). Also, workers who never/occasionally visited the hospital canteen had a lower seroprevalence compared to those with weekly/daily visits (1.9% vs. 3.5%, p=0.004) (**Figure 2, panel D**). This effect was consistent across institutions and professions (**Table S1**).

**Figure 2.**
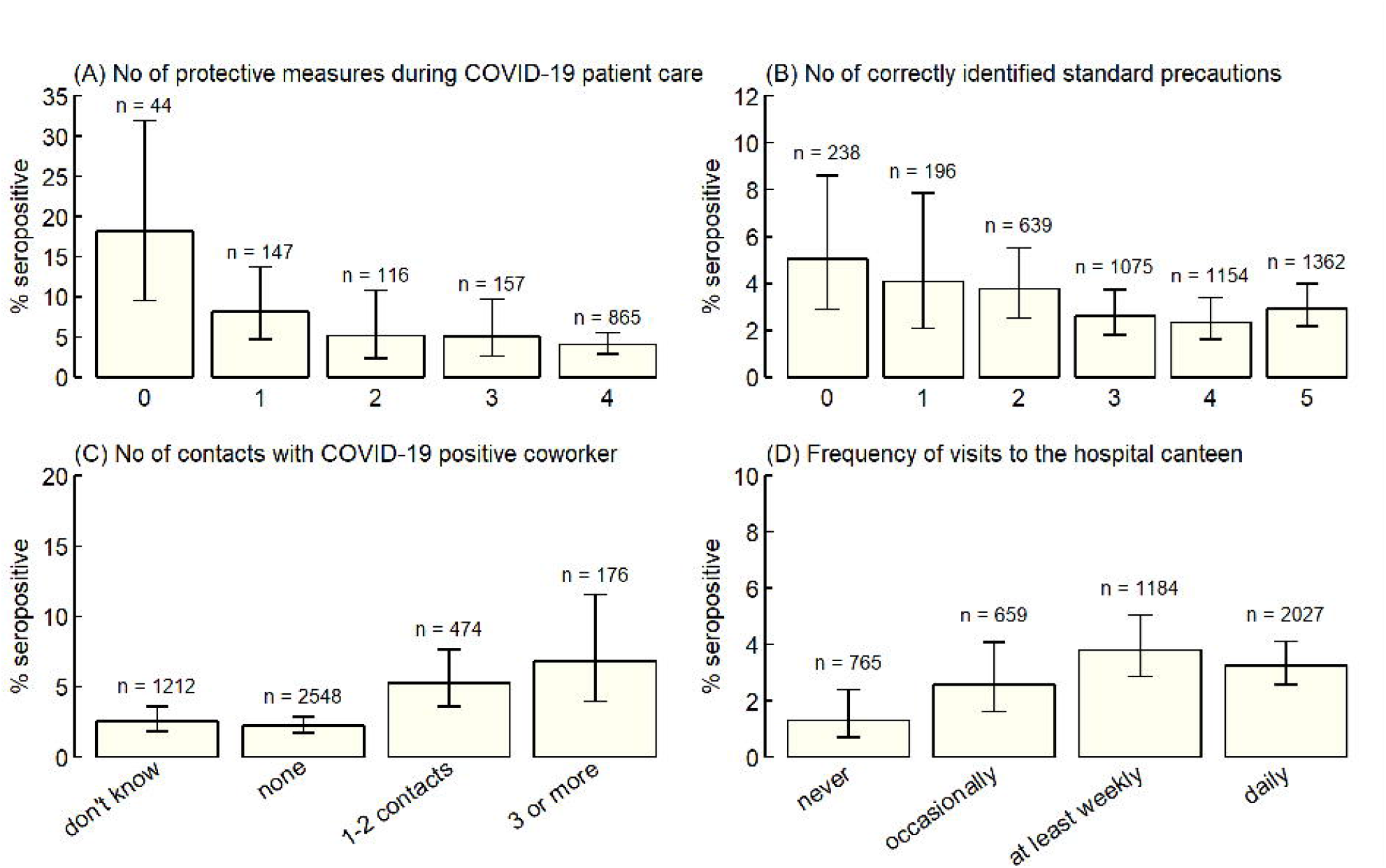
Figure shows (A) number of protective measures used (among face mask, gown, gloves, goggles) while caring for COVID-19 patients; (B) number of correctly identified elements of standard precautions (among hand hygiene, cough etiquette, mask in case of respiratory symptoms, vaccinations, donning of gowns if potential contact with body fluids); (C) number of contacts with COVID-19 positive co-workers; (D) frequency of meals in the hospital canteen.

### Multivariable analyses

In multivariable analysis, exposure to a COVID-19 positive household member remained the strongest risk factor for seropositivity with an adjusted odds ratio (aOR) of 54 (95% CI 31-97) (**Figure 3, Table S2**). Stay in a COVID-19 hotspot was associated with increased risk (aOR 2.2, 95% CI 1.1-3.9), whereas blood group 0 (aOR 0.4, 95% CI 0.3-0.7), active smoking (aOR 0.5, 95% CI 0.3-0.9) and living with children <12 years (aOR 0.3, 95% CI 0.2-0.6) were all associated with decreased risk after correcting for multiple confounder variables.

**Figure 3.**
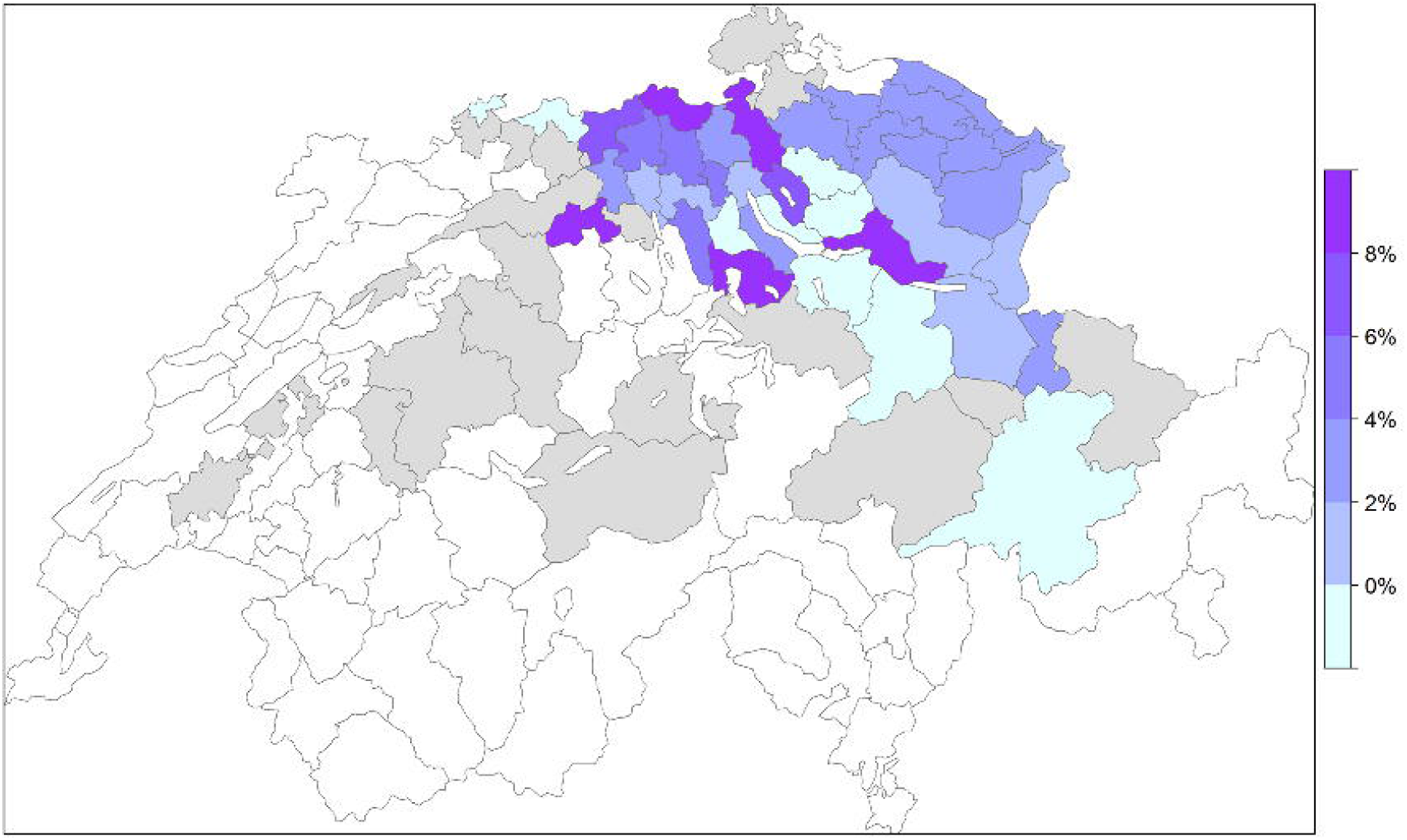
Forest plot showing independent association of baseline, occupational and non-occupational risk factors with seropositivity based on multivariable logistic regression analysis.

Significant occupational factors included close contact with a COVID-19 patient (aOR 2.8, 95% CI 1.5-5.5), exposure to a COVID-19 positive co-worker (aOR 2.0, 95% CI 1.2-3.1), poor knowledge of standard precautions (aOR 2.0, 95% CI 1.3-3.2), as well as having weekly/daily (vs. rarely/never) meals in the hospital canteen (aOR 1.9, 95% CI 1.2-3.1). Both models in the sensitivity analysis did not show any relevant impact of geographic region or institution on the significance level of the variables in the original model (**Table S2**).

## DISCUSSION

In this cross-sectional study of a sample of 4’664 Swiss HCW, 3% of participants had specific SARS-CoV-2 antibodies. Our main findings are that exposure to a COVID-19 positive household member is the strongest risk factor for seropositivity, and that living with children under the age of 12, even after correction for multiple confounders, is clearly associated with decreased risk. Furthermore, we identified several exposures associated with seropositivity which might serve as leverage to further decrease the risk of SARS-CoV-2 acquisition among HCW.

We confirm findings from other studies showing that COVID-19 positive household contacts are the main source of SARS-CoV-2 infection for HCW [8,9]. Our findings are also in line with a Dutch study which concluded that nosocomial transmissions seemed rather uncommon and that multiple hospital introductions from the community are probably responsible for the large part of COVID-19 cases among patients and HCW, at least in a low prevalence setting [10]. Of course, this association might be overestimated given that the directionality of virus transmission cannot be definitely assessed with our study design.

An important finding of our study is that participants living with children <12 years were less likely to be seropositive. A study among over 300’000 HCW households from Scotland has recently found a similar association [11]. In contrast to the Scottish study, we corrected our result for important confounders, including HCW age, full-time working, and leisure activities.

Similary, a population-based cohort from England using large public health databases with over 9 million adults showed that living with children <12 years was not associated with increased risk for SARS-CoV-2 infection, as opposed to living with children aged 12-18 [12]. An intriguing hypothesis for this finding is that certain childhood infections, particularly those with endemic coronaviruses such as HCoV-OC43 or HCoV-HKU1, might confer partial immunity (i.e. cross-immunity) to SARS-CoV-2. In line with this hypothesis, adults aged 15 to 44 years (having presumably an increased probability of living with young children) have been shown to have higher antibody titers against the HCoV-OC43 N protein than older adults [13]. Also, supporting the notion of a rather immunological than a purely epidemiological phenomenon, a German study among over 4’000 COVID-19 patients suggested a less complicated disease course for those with frequent contact to children [14]. This hypothesis was partially confirmed by a recent study that demonstrated pre-existing humoral immunity (including neutralizing antibodies) to be particularly prevalent in children and adolescents [15]. This should be confirmed in prospective studies which evaluate the protective role of humoral and cellular immunity against endemic coronaviruses regarding SARS-CoV-2 acquisition.

Interestingly, stay in an Austrian ski resort where at least one COVID-19 superspreading event had occurred in February/March 2020 was an independent risk factor for seropositivity [16]. Several studies have by now identified an association between the AB0 blood group system and acquisition of COVID-19. Consistently, blood group 0 is considered to have a protective effect as shown in our study, whereas people with a non-0 blood group (mostly A) seem to carry an increased risk [17]. Whether the blood group also determines the course of the disease is less clear [18]. We also observed a lower seroprevalence among active smokers, confirming findings of a meta-analysis [19].

An important question is whether HCW caring for COVID-19 patients are in fact at increased risk for acquiring the disease themselves. A recent meta-analysis concluded that HCW do indeed have an increased risk compared to the general population [20]. Also, frontline HCW in Denmark showed higher seroprevalences than other HCW [21]. Our study confirms these findings, at least for those with close contact to COVID-19 patients. As opposed to other studies [20], a lower level of protection was not significantly associated with seropositivity in multivariable analysis, probably because of our restrictive definition of low protection. Due to the cross-sectional study design we cannot draw valid conclusions regarding the individual benefit of single protective measures such as gloves, gowns or goggles. However, participants performing AGPs as well as those working in intensive care or emergency rooms did not have an increased risk for COVID-19, suggesting that current safety measures are sufficient for these high-risk HCW. Of note, poor knowledge of standard hygiene precautions was associated with detection of SARS-CoV-2 antibodies, supporting efforts to continuously educate HCW regarding basic infection prevention concepts.

Exposure to ill co-workers is a known risk factor for respiratory illness in HCW, not only for COVID-19 but also for other respiratory viral diseases [22]. Across all participating institutions, we identified visits to the hospital canteen as potential risk factor for seropositivity. We found one other study which reported staying in the same HCW break room and eating in proximity to other HCW as risk factor for SARS-CoV-2 transmission [23]. Visiting restaurants other than hospital canteens has previously been shown to be potentially associated with higher risk of SARS-CoV-2 acquisition [24–26]; however, this was not the case in our data. This discrepancy could be explained by the fact that i) the visitor turnover of hospital canteens is much higher than in other eating places and ii) that the probability of a HCW being infectious is higher than for an average visitor to other restaurants. We therefore suggest that hospitals should revisit and potentially reinforce the safety concepts of their canteens and food courts.

Our study has several limitations. First, causality cannot be inferred between exposures and seropositivity. Second, sampling bias may have arisen given that study participation was non-mandatory. Third, we relied on mostly self-reported data in our questionnaire which is subject to recall and other bias. Fourth, because of the low disease prevalence we have to assume that a certain proportion of our serology results are false positive. However, the reported specificity of >99% for the ECLIA [27], the positive confirmatory results in our tested subsample and the overall low proportion of discordant results between PCR and serology supports the validity of our testing approach. Strengths of the study are its large sample size, the inclusion of different types of healthcare institutions across a large geographic area, and consideration of not only occupational but a broad range of non-occupational risk factors. In particular the latter differentiates our study from most other seroprevalence studies performed among HCW.

To conclude, having a COVID-19 positive household member was by far the strongest predictor for SARS-CoV-2 seropositivity among our HCW. Furthermore, we identified several modifiable variables associated with seropositivity, including contact to COVID-19 co-workers, poor knowledge of standard hygiene precautions, and possibly frequent visits to the hospital canteen. Living with children below 12 years of age in the same household was independently associated with decreased risk, an extraordinary finding suggesting an increased role of cross-immunity.

## Data Availability

The authors may provide the original data upon reasonal request.

## Acknowledgments

We would like to warmly thank the large number of employees of the participating health care institutions who either took part in this study themselves or supported it. Furthermore, we thank the laboratory staff for shipment, handling and analysis of the blood samples. In particular, we acknowledge the organizational core team Simone Kessler and Susanne Nigg, who kept all strings between the participating centers and the laboratory and without whom this study would not have been possible.

## Authors contributions

All authors contributed to the conceptualization of the study. CRK and PK supervised the study. CRK, OL, TE, PV and PK were responsible for data curation. TE was responsible for project administration. RP, JS, DF, AB, EL, CM, PR, RS, DV, BW, UB, LR and AF contributed to the investigations. LR provided laboratory resources. SG was responsible for the formal analysis and data visualizations. CRK, PV, and PK were responsible for funding acquisition. CRK, RP and PK wrote the original draft, which was critically reviewed and edited by all authors.

