## Supplemental Table S1 and S2 for "Non-occupational and occupational factors associated with specific SARS-CoV-2 antibodies among Hospital Workers – a multicentre cross-sectional study" (from another version of this preprint: version 2, 2020-11-16)

**SUPPLEMENTS**

Table S1. SARS-CoV-2 seroprevalence between those with frequent and non-frequent visits to the staff restaurant (SR), by profession and institution.

|  | **n participants** | | **% of participants** | **n (seroprevalence) per group** | |
| --- | --- | --- | --- | --- | --- |
|  | rarely / never | weekly to > daily | weekly to > daily | rarely / never | weekly to > daily |
|  | in SR | in SR | in SR | in SR | in SR |
| **Professions** |  |  |  |  |  |
| Other | 167 | 243 | 59% | 0 (0·0%) | 5 (2·1%) |
| Physicians | 106 | 670 | 86% | 0 (0·0%) | 8 (1·2%) |
| Ergotherapists | 18 | 28 | 61% | 0 (0·0%) | 1 (3·6%) |
| Nutritionist | 2 | 16 | 89% | 0 (0·0%) | 1 (6·2%) |
| Research assistant | 6 | 21 | 78% | 0 (0·0%) | 1 (4·8%) |
| Restaurant staff | 15 | 64 | 81% | 0 (0·0%) | 2 (3·1%) |
| Information technology | 14 | 34 | 71% | 0 (0·0%) | 1 (2·9%) |
| Canteen personnel | 1 | 10 | 91% | 0 (0·0%) | 0 (0·0%) |
| Lab-personell | 22 | 32 | 59% | 0 (0·0%) | 0 (0·0%) |
| Speech therapists | 4 | 23 | 85% | 0 (0·0%) | 1 (4·3%) |
| Radiology assistant | 20 | 111 | 85% | 0 (0·0%) | 5 (4·5%) |
| Nurses | 821 | 1305 | 61% | 20 (2·4%) | 63 (4·8%) |
| Physiotherapist | 29 | 106 | 79% | 0 (0·0%) | 6 (5·7%) |
| Facility management | 1 | 17 | 94% | 1 (100·0%) | 1 (5·9%) |
| Secretary | 81 | 181 | 69% | 3 (3·7%) | 3 (1·7%) |
| Social workers | 9 | 27 | 75% | 0 (0·0%) | 0 (0·0%) |
| Technical service | 3 | 37 | 92% | 1 (33·3%) | 2 (5·4%) |
| Administration | 58 | 152 | 72% | 0 (0·0%) | 2 (1·3%) |
| **Institutions^1^** |  |  |  |  |  |
| A | 32 | 76 | 70% | 0 (0·0%) | 1 (1·3%) |
| B | 22 | 87 | 80% | 0 (0·0%) | 3 (3·4%) |
| C | 25 | 84 | 77% | 0 (0·0%) | 3 (3·6%) |
| D | 25 | 88 | 78% | 0 (0·0%) | 0 (0·0%) |
| E | 10 | 107 | 91% | 0 (0·0%) | 4 (3·7%) |
| F | 37 | 81 | 69% | 0 (0·0%) | 1 (1·2%) |
| G | 11 | 112 | 91% | 0 (0·0%) | 3 (2·7%) |
| H | 26 | 133 | 84% | 0 (0·0%) | 4 (3·0%) |
| I | 84 | 84 | 50% | 1 (1·2%) | 6 (7·1%) |
| J | 59 | 110 | 65% | 2 (3·4%) | 5 (4·5%) |
| K | 72 | 99 | 58% | 1 (1·4%) | 2 (2·0%) |
| L | 52 | 138 | 73% | 0 (0·0%) | 1 (0·7%) |
| M | 106 | 211 | 67% | 4 (3·8%) | 1 (0·5%) |
| N | 176 | 272 | 61% | 4 (2·3%) | 14 (5·1%) |
| O | 280 | 563 | 67% | 6 (2·1%) | 26 (4·6%) |
| P | 246 | 777 | 76% | 7 (2·8%) | 32 (4·1%) |

^1^ One institution not shown because of zero seropositives

Table S2. Results of sensitivity analyses, showing multivariable models without region/institution, with region/institution as fixed effects, and with region/institution as random effects (red=positively associated; blue=negatively associated with seropositivity).

|  | **Without inclusion of canton or institution** | | | **Canton and institution as fixed effects** | | | **Canton and institution as random effects** | | |
| --- | --- | --- | --- | --- | --- | --- | --- | --- | --- |
|  | OR | 95% CI | p-value | OR | 95% CI | p-value | OR | 95% CI | p-value |
| Sex (male) | 1·3 | 0·8 - 2·1 | 0·276 | 1·3 | 0·8 - 2·2 | 0·247 | 1·3 | 0·8 - 2·1 | 0·285 |
| Age above 50 | 0·8 | 0·5 - 1·3 | 0·408 | 0·8 | 0·5 - 1·4 | 0·462 | 0·8 | 0·5 - 1·4 | 0·450 |
| BMI above 30 | 1·3 | 0·7 - 2·3 | 0·413 | 1·3 | 0·7 - 2·5 | 0·358 | 1·3 | 0·7 - 2·5 | 0·392 |
| Blood group 0 | 0·4 | 0·3 - 0·7 | 0·001 | 0·4 | 0·2 - 0·7 | 0·001 | 0·4 | 0·3 - 0·7 | 0·001 |
| Smoking | 0·5 | 0·3 - 0·9 | 0·026 | 0·5 | 0·2 - 0·9 | 0·021 | 0·5 | 0·3 - 0·9 | 0·026 |
| Frequent respiratory tract infections | 1·6 | 1 - 2·5 | 0·045 | 1·5 | 0·9 - 2·4 | 0·075 | 1·6 | 1 - 2·4 | 0·058 |
| COVID-19 positive household | 54·2 | 30·9 - 96·5 | < 0·001 | 56·6 | 31·6 - 103·7 | < 0·001 | 54·3 | 30·7 - 96·1 | < 0·001 |
| Symptomatic household member | 2·3 | 1·6 - 3·5 | < 0·001 | 2·4 | 1·6 - 3·7 | < 0·001 | 2·3 | 1·6 - 3·5 | < 0·001 |
| Child(ren) in household | 0·3 | 0·2 - 0·6 | 0·001 | 0·3 | 0·2 - 0·6 | 0·001 | 0·3 | 0·2 - 0·6 | 0·001 |
| Frequent shopping | 0·7 | 0·4 - 1·1 | 0·151 | 0·7 | 0·4 - 1·1 | 0·161 | 0·7 | 0·4 - 1·1 | 0·144 |
| Several leisure activities (now) | 1·2 | 0·8 - 1·8 | 0·365 | 1·3 | 0·9 - 1·9 | 0·235 | 1·2 | 0·8 - 1·8 | 0·308 |
| Stay in COVID-19 hotspot | 2·2 | 1·1 - 3·9 | 0·013 | 2·7 | 1·4 - 4·9 | 0·002 | 2·3 | 1·2 - 4·3 | 0·008 |
| High work percentage | 0·9 | 0·6 - 1·4 | 0·536 | 0·9 | 0·6 - 1·4 | 0·591 | 0·9 | 0·6 - 1·4 | 0·552 |
| Patient contact | 1·3 | 0·6 - 2·6 | 0·538 | 1·5 | 0·7 - 3·1 | 0·322 | 1·3 | 0·6 - 2·8 | 0·441 |
| Close patient contact | 1·1 | 0·6 - 2·2 | 0·721 | 1·1 | 0·6 - 2·1 | 0·863 | 1·1 | 0·6 - 2·1 | 0·738 |
| Aerosol generating procedures | 0·7 | 0·5 - 1·1 | 0·150 | 0·6 | 0·4 - 1 | 0·068 | 0·7 | 0·4 - 1·1 | 0·113 |
| COVID-19 patient contact | 0·7 | 0·3 - 1·4 | 0·284 | 0·6 | 0·3 - 1·3 | 0·222 | 0·7 | 0·3 - 1·4 | 0·272 |
| Close COVID patient contact | 2·8 | 1·5 - 5·5 | 0·002 | 3·1 | 1·6 - 6·2 | 0·001 | 2·8 | 1·5 - 5·5 | 0·002 |
| Low protection with COVID patient | 1·7 | 1 - 2·8 | 0·062 | 1·6 | 0·9 - 2·8 | 0·099 | 1·6 | 1 - 2·8 | 0·072 |
| COVID-positive coworker | 2 | 1·2 - 3·1 | 0·004 | 1·9 | 1·2 - 3 | 0·009 | 1·9 | 1·2 - 3·1 | 0·006 |
| Poor knowledge of hygiene standards | 2 | 1·3 - 3·2 | 0·002 | 2·2 | 1·4 - 3·5 | 0·001 | 2·1 | 1·3 - 3·3 | 0·001 |
| Frequent meals in staff canteen | 1·9 | 1·2 - 3·1 | 0·010 | 1·9 | 1·2 - 3·1 | 0·014 | 1·9 | 1·2 - 3·1 | 0·010 |
|  |  |  |  | Chi-squared | df | p-value | sd of random effect | |  |
| Institution | - | - | - | 30·7 | 22 | 0·103 | 0·305 | 23 groups |  |
| Region (geographic clustering) | - | - | - | 2·9 | 6 | 0·824 | 0·001 | 7 groups |  |
